# Genetically Determined Blood Pressure, Antihypertensive Medications, and Risk of Intracranial Aneurysms and Aneurysmal Subarachnoid Hemorrhage: A Mendelian Randomization Study

**DOI:** 10.1101/2023.02.16.23286069

**Authors:** Hanchen Liu, Huiqin Zuo, Ospel Johanna, Rui Zhao, Pengfei Yang, Qiang Li, Xiaolei Lin, Yu Zhou, Jianmin Liu

## Abstract

**Background:** Observational studies suggest that different classes of antihypertensive drugs may have different effects on the occurrence of intracranial aneurysm(IA) and subarachnoid hemorrhage(SAH). However, the reported effects in previous studies are inconsistent, and randomised data are absent. We performed a two-sample Mendelian randomization(MR) analysis to study the causal effects of genetically determined blood pressure(BP) and genetic proxies for antihypertensive drug classes on the risk of IA and SAH.

**Methods:** Genetic instruments and outcome data were obtained from independent genome-wide association studies(GWAS) or published data, which were exclusively restricted to european ancestry. Causal relationships were identified using inverse-variance weighted MR analyses and a series of statistical sensitivity analyses. The Finngen consortium was used for repeated analysis to verify results obtained from above GWAS.

**Results:** Two-sample MR analysis showed that genetically determined Systolic BP, Dystolic BP and Pulse Pressure were related to higher risk of IA and SAH. Based on indentified single nucleotide polymorphisms (SNPs) that influence the effect of calcium channel blockers (CCB, 43 SNPs), beta-blockers (BB, 30 SNPs), angiotensin-converting enzyme inhibitors (ACEI, 16 SNPs), angiotensin receptor blockers (ARB, 11 SNPs), and thiazides (5 SNPs), genetically determined effect of CCBs was associated with higher risk of IA (OR, 1.08 [95%CI, 1.04-1.12], P=1.21×10 ^-5^) and SAH (OR, 1.07 [95%CI, 1.02-1.12], P=2.05×10 ^-3^). No associations were found between other antihypertensive drugs and risk of IA or SAH. The effect of CCBs on SAH was confirmed in Finngen consortium samples(OR, 1.04 [95%CI, 1.00-1.08], P=0.042).

**Conclusion:** This MR analysis supports the protection effect of hypertension control on the occurrence of intracranial aneurysm and subarachnoid hemorrhage. However, genetic proxies for calcium channel blockers were associated with increased risk of intracranial aneurysm and subarachnoid hemorrhage. Further studies are required to confirm these findings and investigate the underlying mechanisms.

## Introduction

Hypertension is a common comorbidity of patients with intracranial aneurysms and may cause de novo aneurysms to appear and existing ones to grow.^1, 2^ It is thought that adequate antihypertensive treatment to normalize blood pressure could significantly decrease aneurysm rupture risk. ^3^ Some investigators further evaluated the effect of various kind of antihypertensive drugs (calcium channel blockers [CCB],beta-blockers [BB], angiotensin-converting enzyme inhibitors [ACEI], angiotensin receptor blockers[ARB], or thiazide diuretics)on the incidence of IA or SAH, and found different effects depending on the type of drugs. However, the literature is inconclusive in this regard. For example, while some studies suggest that ARBs and ACEI are effective in preventing aneurysm rupture,^4^ others found no such effect.^5^ One cross-sectional study even found CCBs were independently associated with the presence of IAs.^6^

Mendelian Randomization(MR) analysis is a complementary approach for assessing the causal effects of risk factors in observational studies using genetic variation.^7^ It uses genetic variants (single-nucleotide polymorphisms [SNPs]) as proxies for exposure to certain risk factors to investigate their effects on an outcome of interest. Due to the random allocation of SNP alleles at conception, MR offers an opportunity to overcome limitations inherent to traditional observational studies, such as residual confounding and reverse causation. In addition, naturally occurring variation in genes encoding drug targets could also be used as proxies for medications targeting these targets to examine the effect of their therapeutic effect on disease outcomes. Such a drug-target mendelian randomization can be used to mimic pharmacological clinical trials and has been used previously to anticipate clinical benefits and adverse effects of therapeutic interventions.^11^

Herein, we performed a two-sample MR analysis to investigate the causal effects of genetically determined BP and genetic proxies for antihypertensive drug classes on the risk of IA and SAH.

## Methods

### Study Design and Data Availability

Two sample Mendelian randomisation—using two different study samples to estimate the instrument-risk factor and instrument-outcome associations to estimate a causal effect of the risk factor on the outcome, was performed in this study.Summary statistics were obtained from published and publicly available GWAS studies were used in this study (**Table 1**). This study was conducted and reported in accordance with the guidelines for Strengthening the Reporting of Observational Studies in Epidemiology– Mendelian randomization (STROBE-MR).^8^

**Table 1.** Characteristics of the Used GWAS

| **Variable** | **First author (year)** | **Consortium** | **Sample size** | **Ancestry** | **Sex** | **Unit** |
| --- | --- | --- | --- | --- | --- | --- |
| Diastolic blood pressure | Evangelou, E(2018) | International Consortium of Blood Pressure | 757,601 | European | Males and Females | 1 mmHg |
| systolic blood pressure | Evangelou, E(2018) | International Consortium of Blood Pressure | 757,601 | European | Males and Females | 1 mmHg |
| Pulse pressure | Evangelou, E(2018) | International Consortium of Blood Pressure | 757,601 | European | Males and Females | 1 mmHg |
| subarachnoid hemorrhage | FinnGen biobank (2021 ) | NA | 205,196 | European | Males and Females | NA |

### Instrumental variables for exposure

Genetic instruments for Systolic Blood Pressure(SBP), Dystolic(DBP) and Pulse Pressure(PP) were obtained from the summary statistics of the GWAS meta-analysis consisting of 757,601 individuals (458,577 from UK Biobank and 299,024 from the International Consortium of Blood Pressure) of European ancestry.^9^ A total of 458, 457 and 332 independent SNPs were associated with SBP, DBP and PP respectively at the genome-wide significant level (P < 5 × 10 ^− 8^, **Table S1-S3**).

Five commonly used antihypertensive drugs were selected, including ACEI, ARB, BB, CCB and thiazide diuretics, according to the latest guidelines.^10^ Genetic variants in the drug targeted genes associated with BP at genome-wide signicant level ^11-15^ were used as instruments for the effects of antihypertensive drugs. Specifically, genes encoding the pharmacologic targets related to the effect of common antihypertensive drugs on BP were identified in DrugBank (https://www.drugbank.ca/).^16^ SNPs corresponding to the functional genes as well as their promoter and enhancer regions were screened in GeneCards (https://www.genecards.org/).^17^ Finally, variants associated with SBP at the genome-wide significant level (P< 5 × 10 ^− 8^) were clumped to a linkage disequilibrium (LD) threshold of r^2^ < 0.4 using the 1000G European reference panel as candidate instrument for each antihypertensive drug class. This relatively lenient LD threshold allows for an increase in proportion of variance explained and thus in statistical power.^18,19^

### Outcome Data

Summary-level outcome data were obtained from the GWAS on IA and SAH, which consisted of 23 cohorts with a total of 7, 495 cases and 71, 934 controls of European ancestry.^20^ We used the summary statistics excluding the UK Biobank data as outcomes to avoid the risk of bias due to sample overlap.^21^

Additionally, summary genetic association data were obtained from 2127 SAH cases and 203,068 controls of European ancestry from the Finngen consortium for repeated MR analysis to verify the results obtained from the above GWAS about the association between antihypertensive drugs and SAH. Analysis of the association between antihypertensive drugs and IA was not repeated due to lack of another independent GWAS data.

### Statistical Analysis

The primary MR analysis was conducted using the inverse variance weighted (IVW) MR method. In addition to the three pre-requisites required for all types of MR analyses (1: strong association between genetic instrument and the exposure, 2: no confounding between the genetic instrument and outcome, and 3: entire association between genetic instruments and the outcome explained by the exposure),^22^ IVW MR additionally assumes that all genetic variants are valid instruments and combines the SNP-specific Wald estimates 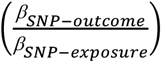 using inverse of the corresponding variance.^23^ A series of sensitivity analyses was conducted to evaluate the robustness of our results. First, Cochran’s Q statistic was used to assess the heterogeneity between individual genetic variants in the IVW MR method. In case of heterogeneity, results from the weighted median method were adopted, which can provide consistent effect estimates even when more than 50% of the information comes from invalid or weak SNPs.^24^ Next, horizontal pleiotropy was assessed using MR-Egger and controlled using MR pleiotropy residual sum and outlier (MR-PRESSO) if directional pleiotropy existed. Finally, leave-one-SNP-out analysis was performed, in which SNPs were systematically removed, to assess if results were driven by single SNP.

Results were presented as odds ratios (OR) and 95% confidence intervals (CIs) for IA and SAH per genetically predicted unit log-transformed increase in each trait. Association were considered significant after correcting for multiple testing across three BP indexes and five classes of antihypertensive drugs using the Bonferroni method (P=0.05/8=0.00625). All analyses were performed via TwoSampleMR (version 0.5.6), Mendelian randomization (version 0.5.1), and MRPRESSO (version 1.0) packages in R version 4.2.2. ^25^

## Results

### Genetically determined blood pressure and risk of IA/SAH

As shown in **Figure 1**, MR analyses showed statistically significant associations between genetically determined SBP, DBP, PP and risk of IA. For every 10 mm Hg/5 mm Hg/1 mm Hg increase in genetically determined SBP/DBP/PP, the risk of IA increased by 67% (OR, 1.67 [95%CI, 1.45-1.92], P=8.72×10 ^-13^), 64% (OR, 1.64 [95%CI, 1.84-3.31], P=6.20×10 ^-20^), and 6% (OR, 1.06 [95%CI, 1.03-1.08, P=6.01×10 ^-8^), respectively. In addition, we also observed a significant increase in the risk of SAH per 10 mm Hg/5 mm Hg/1 mm Hg increase in genetically determined SBP (OR, 1.82 [95%CI, 1.53-2.18], P=1.94×10 ^-11^), DBP (OR, 1.67 [95%CI, 1.46-1.91], P=4.97×10 ^-14^) and PP (OR, 1.07 [95%CI, 1.05-1.09], P=7.84×10 ^-13^).

**Figure 1.**
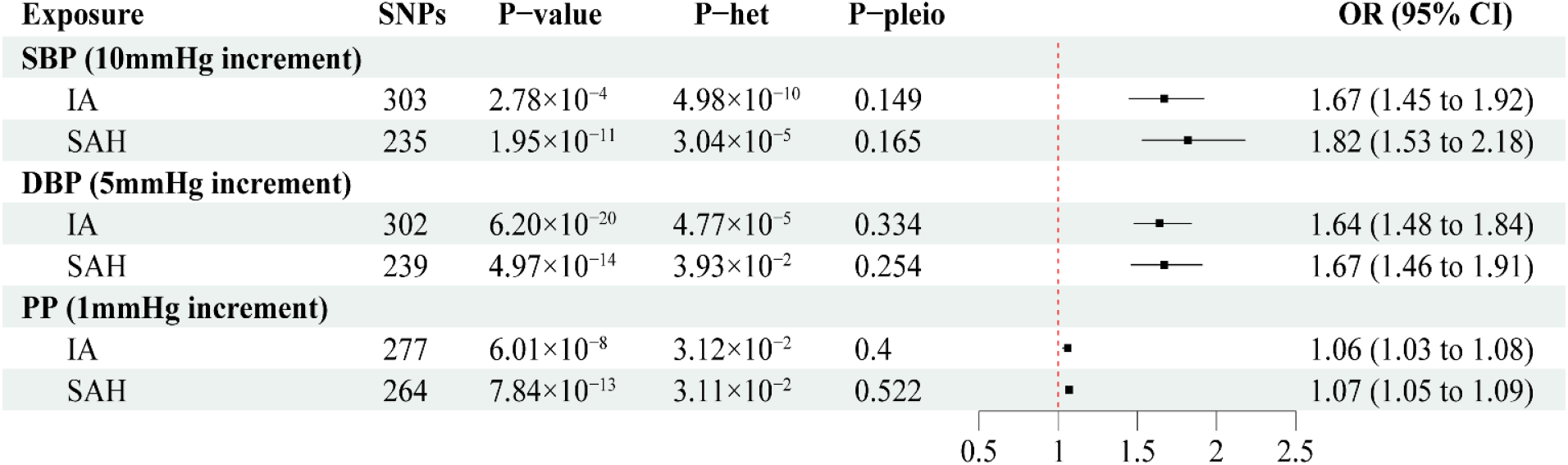
MR associations between genetically determined blood pressure and the risk of Intracranial aneurysm development and Subarachnoid Hemorrhage Genome-wide significantly associated (P < 5 × 10 ^− 8^) independent (LD R^2^ = 0.001, clumping distance = 10,000 kb) SNPs were used as instruments. MR, Mendelian randomization; SNP, single nucleotide polymorphism; OR, odds ratio; CI, confidence interval; SBP, systolic blood pressure; DBP, diastolic blood pressure; PP, pulse pressure; IA, Intracranial aneurysm; SAH, Subarachnoid Hemorrhage; P-het,P value in the Q statistic for heterogeneity; P-pleio, P value in the Egger intercept. P value in the Q statistic for heterogeneity; P-pleio, P value in the Egger intercept.

In sensitivity analyses, Cochran’s Q statistic suggested potential heterogeneity among individual SNP in IVW MR. Therefore, we refer to the weighted median method in the presence of heterogeneity, which showed similar results as IVW MR (**Table S5**). Leave-one-SNP-out analysis showed that results were robust to all SNPs and were not driven by any single SNP (**Figure S1**).

### Genetically determined effects of antihypertensive drugs and risk of IA/SAH

A total of 16, 11, 30, 43 and 5 independent SNPs were asscociated the antihypertensive effect of ACEI, ARB, BB, CCB and thiazide diuretics at the genome-wide significant level (**Table S4**). As shown in **Figure 2**, genetically determined effect of CCB was positively associated with higher risk of IA (OR, 1.08 [95%CI, 1.04-1.12], P=1.21×10 ^-5^) and SAH (OR, 1.07 [95%CI, 1.02-1.12], P=2.05×10 ^-3^). However, we did not find any significant associations between other antihypertensive drugs and risk of IA or SAH (**Table S6**). Sensitivity analysis using the Cochran’s Q statistic indicated no notable heterogeneity and directional pleiotropy across instrument SNP effects (**Table S6**). There was no distortion in the leave-one-SNP-out plot, suggesting that no single SNP was driving the observed effect in the analysis (**Figure S1**).

**Figure 2.**
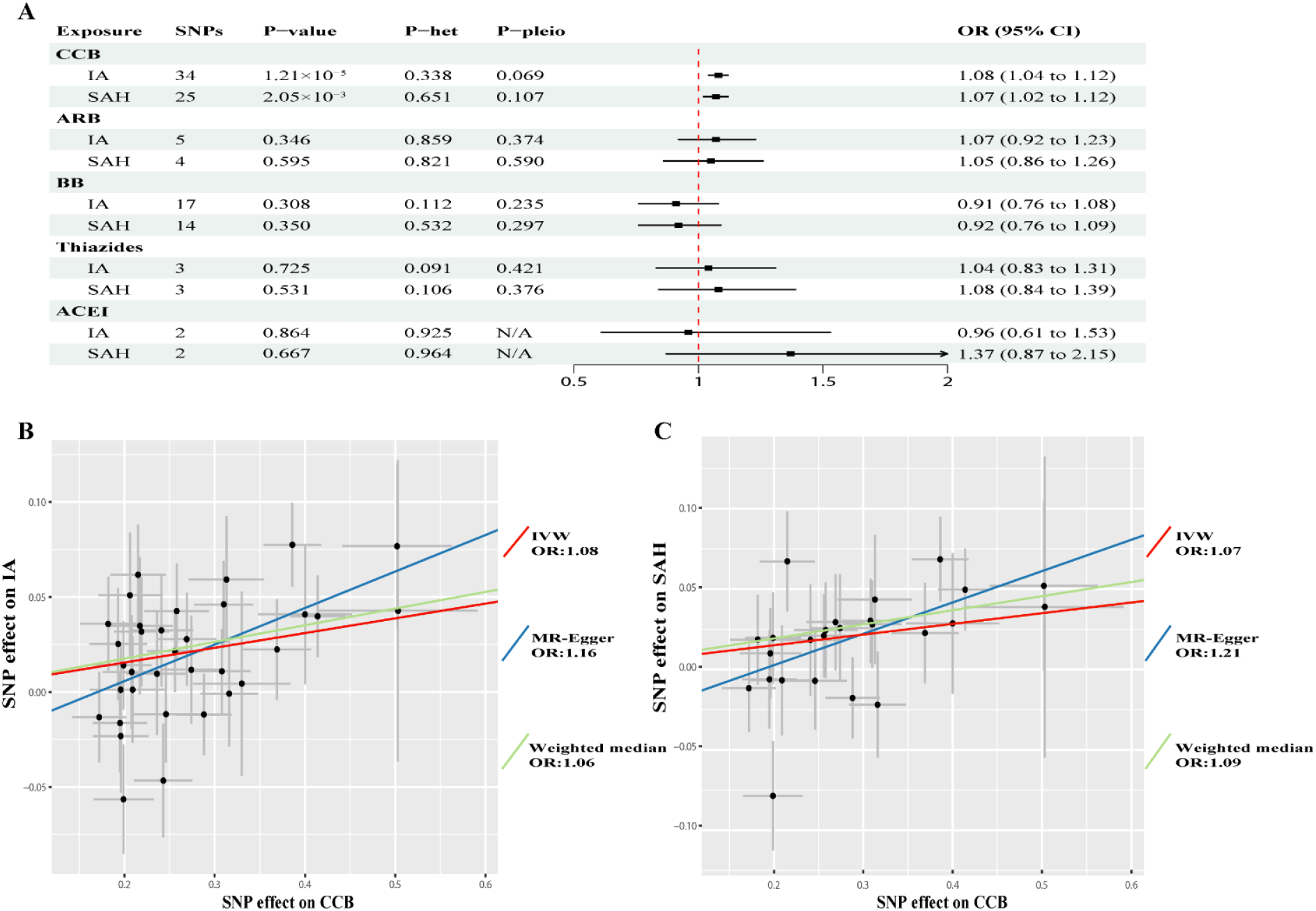
MR associations between genetically determined antihypertensive drugs and the risk of Intracranial aneurysm development and Subarachnoid Hemorrhage **A**.Inverse-variance weighted(IVW) estimates for the association between a genetically determined unit increase in exposure on the risk of intracranial aneurysm(IA) and subarachnoid hemorrhage(SAH). **B** and **C**, Scatter plots of individual single-nucleotide polymorphisms (SNPs) effects and estimates from different Mendelian randomization (MR) methods for the effect of (**B**) calcium channel blockers (CCB) on IA and (**C**) CCB on SAH. P-het is the P value belonging to the Q statistic for heterogeneity. P-pleio is the P value belonging to the Egger intercept. MR, Mendelian randomization; SNP, single nucleotide polymorphism; OR, odds ratio; CI, confidence interval; ACEI: angiotensin-converting enzyme inhibitor; ARB: angiotensin receptor blocker; CCB: calcium channel blockers; BB: beta-blockers; ACEI: angiotensin-converting enzyme inhibitors; ARB: angiotensin receptor blockers; IA, Intracranial aneurysm; SAH, Subarachnoid Hemorrhage; P-het,P value in the Q statistic for heterogeneity; P-pleio, P value in the Egger intercept.

### Replication of findings

Repeated analyses using another SAH GWAS sample of European ancestry (the Finngen consortium) yielded similar results. An 47%, 31% and 3% increase in SAH risk per 10 mm Hg/5 mm Hg/1 mm Hg increment was observed in genetically determined SBP (OR, 1.47 [95%CI, 1.30-1.66], P=8.63×10 ^-10^), DBP (OR, 1.31 [95%CI, 1.18-1.45], P=1.54×10 ^-7^) and PP (OR, 1.03 [95%CI, 1.01-1.06], P=1.15×10 ^-3^), respectively. In addition, genetically determined effect of CCB also showed positive association with risk of SAH (OR, 1.04 [95%CI, 1.00-1.08], P=0.042)(**Table S7**). Sensitivity analysis revealed no heterogeneity and pleiotropy in these analysis.

## Discussion

Using large-scale genetic data from GWAS, we confirmed the deleterious effect of hypertension on the occurrence of IA and SAH. Furthermore, we observed that genetically determined CCBs may increase the risk of IA (OR, 1.08 [95%CI, 1.04-1.12], P=1.21×10 ^-5^) and SAH (odds ratio, 1.07 [95%CI, 1.02-1.12], P=2.05×10 ^-3^), which held true in another large GWAS, namely the Finnegan population, as well (OR, 1.04 [95%CI, 1.00-1.08], P=0.042). For other hypertensive drugs, neither harmful nor beneficial effects on IA and SAH were observed.

Consistent with previous studies, our findings provide complementary evidence for the role of hypertension in IA development and SAH occurence, and underscore the importance of blood pressure control for preventing IA and SAH.^26, 27^ For every 10 mmHg increase in genetically determined SBP, the risk of IA and SAH increased by 67% and 82%, while for every 5 mm Hg increase in genetically determined DBP the risk of IA increased by 64% and 67%. Besides SBP and DBP, our study found that PP may also be an important factor contributing to the occurrence of IA and SAH. For every 1 mm Hg increase in genetically determined PP, the risk of IA and SAH increased by 6% and 7%, respectively.

Regarding antiypertensive drugs, their effect on IA and SAH is somewhat controversial in the existing literature, with inconsistent findings among different studies. One case-control study compared an IA cohort (n = 1960) with a matched normal population (n = 1960), and found that CCB were independently associated IA (1.49 [1.11–2.00]). However, another cross-sectional study including 310 patients with ruptured and 887 patients with unruptured IA, CCBs were found to be inversely associated with ruptured IA (OR, 0.41; 95% CI 0.30-0.58). The authors hypothesized that CCB may be effective in preventing unruptured IAs from rupturing. In contrast to this, we found CCB to have deleterious effects on both IA and SAH. These discrepancy may be related to differences in study design, inherent limitations of previous observational studies, or limited follow-up periods.

Assuming that CCBs truly cause deleterious effects on IA and SAH, the underlying mechanisms are still unknown, but the mechanisms may be similar to aortic aneurysms, whose course is also negatively influenced by CCB ^28^ : Not only do Marfan mice treated with CCBs show accelerated aneurysm expansion, rupture, and premature lethality, patients with Marfan-associated and other forms of inherited thoracic aortic aneurysms taking CCBs also show an increased risk of aortic dissection and need for aortic surgery when compared to other antihypertensive agents. The most likely reason is that CCBs enhance ERK1/2 activation, and PKCβ mediates CCB-induced aortic aneurysm exacerbation.^28^ Furthermore, Ca^2+^ channel blockade seems to induce medial smooth muscle cell apoptosis in thoracic aortae of spontaneously hypertensive patients.^29^ Rupture or IA is also thought to be related to smooth muscle cell apoptosis.^30^ Whether the mechanisms observed in thoracic aneurysms can really be translated to IA is not entirely clear. Overall, it seems that in our study, the deleterious effects of CCBs on IA and SAH may outweigh the protective effects caused by blood pressure control. It is certainly premature to say that based on these findings, CCBs should be abandoned, and clearly, further research is needed in this regard. That being said, choosing other anti-hypertensive drugs other than CCBs as first-line agents may be reasonable.

Despite numerous observational studies showing that ARB and ACEI may be protective for aneurysm rupture,^4^ our study failed to confirm these associations. However, our results cannot completely rule out their effect on IA/SAH because the limited number of SNPs in this study may not provide sufficient statistical power.

Our study has some limitations. First, type-I errors may occur due to multiple testing. However, the associations observed persisted even after adjusting for multiple testing. Second, MR analysis explores the effects of lifetime exposure, while drug exposure is typically much shorter, and blood pressure may also have age-dependent effects. The causal effect sizes obtained from our study therefore most likely do not adequately represent the relationship between exposure and outcome.^31^ Therefore, mendelian randomization studies can only provide supplementary evidence, and clearly, further randomized controlled trials are warranted to investigate the effects of drugs on IA and SAH. Third, since our genetic agents were determined according to drug targets, we only focused on the pharmacodynamics of drugs but not on their pharmacokinetics. Therefore, the existing analysis can-not fully reflect the relationship between drugs and outcomes. Lastly, the data we used were all from individuals of European ancestry; thus, the results of this study may not necessarily apply to other populations.

## Data Availability

All data in this study are publicly available

## Conclusion

In this MR analysis, genetically determined hypertension was associated with a higher risk of IA and SAH. Although CCBs have BP lowering properties, they may increase the risk of IA and SAH. As CCBs are widely used in patients with SAH, further (mechanistic) studies are needed to determine the effect of CCBs on IA and SAH.

## Sources of funding

This study was partially supported by grants from the National Natural Science Foundation of China (No. 81400979 and 81870931), the SanHang Program of the Naval Medical University, and the ‘Climbing’ program of Changhai hospital.

## Disclusure

The authors report no conflicts.

## Supplemental Material

Tables S1-S7

Figure S1

STROBE-MR Checklist

## Non-standard Abbreviations and Acronyms

ACEI: angiotensin-converting enzyme inhibitor;
ARB: angiotensin receptor blocker;
IA: intracranial aneurysm;
SAH: subarachnoid hemorrhage;
MR: Mendelian randomization;
GWAS: genome-wide association studies;
SNPs: single nucleotide polymorphisms;
BP: blood pressure;
CCB: calcium channel blockers;
BB: beta-blockers;
ACEI: angiotensin-converting enzyme inhibitors;
ARB: angiotensin receptor blockers;
OR: odds ratios;
CI: confidence intervals;
LD: linkage disequilibrium;
IVW: inverse variance weighted;
DBP: diastolic blood pressure;
SBP: systolic blood pressure;
PP: pulse pressure.

## Notes

### Competing Interest Statement

The authors have declared no competing interest.

